# A Novel Diathesis-Stress Model of Comorbid Early Onset Psychiatric Disorders

**DOI:** 10.1101/2025.04.08.25325481

**Authors:** Charlotte B. Caswell, Niki Hosseini-Kamkar, Sylvia M. Cox, Nicole Palacio Prada, Maisha Iqbal, Maja Nikolic, Tobias Banaschewski, Gareth J. Barker, Arun L.W. Bokde, Rüdiger Brühl, Sylvane Desrivières, Herta Flor, Hugh Garavan, Penny Gowland, Antoine Grigis, Andreas Heinz, Jean-Luc Martinot, Maire-Laure Paillère Martinot, Eric Artiges, Frauke Nees, Dimitri Papadopoulos Orfanos, Luise Poustka, Michael N. Smolka, Sarah Hohmann, Nilakshi Vaidya, Henrik Walter, Robert Whelan, Gunter Schumann, Tomáš Paus, Marco Leyton, IMAGEN Consortium

## Abstract

**Importance:** Psychiatric comorbidity is the norm. Identifying transdiagnostic risk factors will inform our understanding of developmental pathways and early intervention targets.

**Objective:** We recently reported that many psychiatric outcomes are predicted by a three-factor model composed of adolescent externalizing (EXT) behaviors, early life adversity, and dopamine autoreceptor availability. Here, we investigated whether this model could be reproduced in a large population-based sample using functional magnetic resonance imaging (fMRI) instead of positron emission tomography.

**Design:** Data were collected by the IMAGEN consortium beginning in 2010 when cohort members were 14 years old, with follow-up testing at ages 16 and 19. These longitudinal data were used to predict psychiatric disorders by 19 years of age.

**Setting:** Participants were recruited from secondary schools across Europe.

**Participants:** Adolescents (n = 1338) with fMRI, behavioural, diagnostic, and early life trauma data.

**Main Outcomes and Measures:** Binary regression models tested whether a combination of EXT behaviors, childhood trauma, and mesocorticolimbic reward anticipation responses at age 14 or 19 predicted the presence of a disorder by age 19.

**Results:** A total of 1338 participants had the required data (52.4% female). In all models, EXT and adversity scores were significant predictors (*p* < 0.001). Reward anticipation responses in the ventral striatum, caudate, putamen, and anterior cingulate cortex (ACC) at age 14 (*p* ≤ 0.05) and in the ventral striatum at age 19 (*p* ≤ 0.029) were predictors in their respective models. The three-factor models overall were highly significant (*p* < 1.0 x 10^-21^), yielding greater predictive strength than each factor alone. They had an accuracy of nearly 75%, accounting for ≥ 11% (Nagelkerke R^2^) of the variance in psychiatric disorders. The relationship between trauma and diagnoses was partially mediated by higher EXT (indirect path *B* = 0.0535, 95% CI = 0.0301- 0.0835), and moderated by fMRI responses in the ACC (*p* = 0.0038) and putamen (*p* = 0.0135) at age 14.

**Conclusions and Relevance:** The results extend our previous findings, increasing confidence in a novel diathesis-stress model of commonly comorbid early onset psychiatric disorders. The results have implications for diagnostic classification schemes and pleiotropic views of psychiatric disorder etiology.

**Key Points:** *Question:* What factors contribute to a diathesis-stress model of commonly comorbid psychiatric disorders?

*Findings:* A combination of childhood adversity, adolescent externalizing behavior, and lower mesocorticolimbic reward cue reactivity predicted psychopathology by age 19 in a large population-based cohort. Liability was modified by interactions between trauma and mesocorticolimbic responses, with trauma effects larger in those with smaller striatal and anterior cingulate cortex responses.

*Meaning:* The study identifies a transdiagnostic diathesis-stress model of early onset psychiatric disorders.

## INTRODUCTION

Psychiatric comorbidity is the norm (1, 2, 3). The ubiquity of this observation raises the possibility that many psychiatric disorders reflect shared etiologic factors (3, 2). The best-established transdiagnostic risk factor is early life adversity (6, 7, 8, 9, 10, 11, 12). Despite this, even severe adversity is associated with psychiatric disorders in only some people (12, 13, 14).

The varying responses to early adversity (15, 16) might reflect individual differences in temperamental traits (17, 18, 19, 20) and mesocorticolimbic system reactivity (21, 22, 23).

Indeed, both adolescent externalizing (EXT) behaviors and mesocorticolimbic responses to reward-related events predict poor outcomes across a wide range of disorders (25, 26, 27). Despite being widely replicated, these effects are not large when studied in isolation.

Recently, we tested whether psychiatric outcomes are better predicted by combining early life adversity, adolescent EXT, and midbrain dopamine autoreceptor availability, as compared with considering the features independently. In this preliminary work, carried out with positron emission tomography (PET) in young adults followed since their birth (n = 52), the combination of factors identified those with a lifetime disorder with high statistical robustness and accuracy (27). The current study aimed to reproduce this finding in a larger sample (n = 1338) using task-based functional magnetic resonance imaging (fMRI) instead of PET. The use of fMRI allowed us to measure, during adolescence, brain reactivity throughout the mesocorticolimbic pathway with a particular interest in regions implicated in reward processing. The larger sample size provided sufficient statistical power to examine how the three factors affect each other. Together, this study tested a novel diathesis-stress model whereby diathesis, a form of “predispositional” vulnerability, interacts with adversity-induced stress (17).

## METHODS

### Participants

Participants were 1338 youth from the longitudinal IMAGEN project (29). They were included if they had completed the Strengths and Difficulties Questionnaire (SDQ; 30) at 14 and/or 16 years old, the Childhood Trauma Questionnaire (CTQ; 30) at age 19, and the Development and Wellbeing Assessment (DAWBA; 32) at ages 14, 16, and/or 19. An externalizing (EXT) score was derived from three SDQ hyperactivity subscale items (e.g., ‘constantly fidgeting or squirming’) and five conduct subscale items (e.g., ‘often fights with other children or bullies them’; see eMethods in Supplement). Both subscales were converted to a 10-point scale, and their mean was taken for each age. For participants who completed the SDQ at both 14 and 16 years old, the mean was used to represent adolescent EXT. The DAWBA uses six probability bands ranging from <0.1% likelihood to >70% likelihood of having a given disorder in the previous year, confirmed by a clinician (see eMethods in Supplement). Research ethics approval was obtained at each data collection site (<https://www.imagen-project.org/ethics>). For more details about recruitment and questionnaire administration, see < https://www.imagen-project.org/>.

### Functional Magnetic Resonance Imaging

As reported in greater detail elsewhere (29), eight IMAGEN sites used harmonized gradient echo-planar imaging (EPI) sequences to acquire blood oxygen level dependent BOLD data at 3T (Oblique slice plane, repetition time = 2,200 ms, echo time = 30 ms, flip angle = 75°, matrix = 64x64, field of view = 220 x 220 mm^2^, 40 slices with 2.4 mm slice thickness, and 1 mm gap; 300 volumes per run).

### Monetary Incentive Delay Task

A version of the monetary incentive delay (MID) task, originally developed by Knutson et al. (2000), was performed at ages 14 and 19 during fMRI in which rewards were represented by points rather than monetary values (see eMethods in Supplement).

### fMRI analyses

We obtained fMRI data pre-processed by the IMAGEN core sites (previously described in detail, e.g., 34). In brief, pre-processing was conducted in SPM8 (Wellcome Trust Centre for Neuroimaging, London) and included slice-time correction, non-linear warping to the MNI (Montreal Neurological Institute) space, and smoothing with a 5-mm full-width-at-half-maximum (FWHM) isotropic Gaussian kernel.

Three contrasts of the MID task were selected to investigate reward anticipation: ‘large – small win’, ‘large – no win’, ‘small – no win’ (e.g., 35, 23, 36). Region of interest (ROI) analyses were performed using the MarsBar toolbox for SPM12. The ROIs were the midbrain (comprising left and right substantia nigra and ventral tegmental area), the striatum with three sub-regions (ventral striatum {VS}, caudate, putamen), and the cerebral cortex (orbitofrontal cortex {OFC}, ventromedial prefrontal cortex {vmPFC}, and anterior cingulate cortex {ACC}; see eFigure 1 in Supplement). The midbrain ROI was from the Duke Atlas from the Addock laboratory (37).

**Figure 1.**
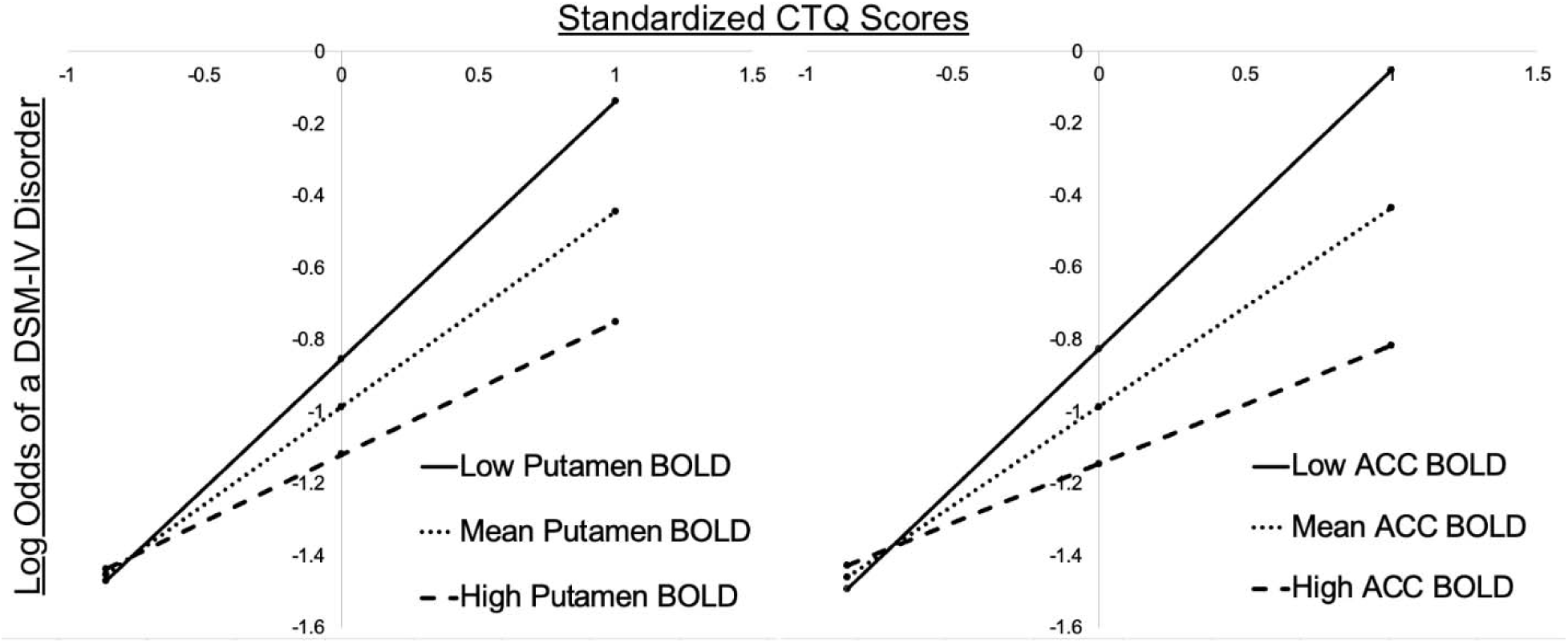
Moderation of Childhood Trauma by Reward Anticipation fMRI Response. Simple slopes plot showing the log odds of DAWBA identified DSM-IV disorders at low, mean, and high standardized levels of CTQ score and low, mean, and high levels age 14 Large - Small reward anticipation activations in the (A) L putamen and (B) ACC.

Bilateral vmPFC and ACC ROIs were based on coordinates from previous IMAGEN reports (38, 39). All other coordinates were selected from a meta-analysis of activations during reward anticipated-related fMRI tasks (40).

### Statistical Analyses

All analyses were performed using SPSS version 29.0.1.1 (IBM Corp). Binary logistic regression models were constructed for each ROI. The model’s three factors were i) BOLD activity for a given ROI, ii) EXT scores, and iii) CTQ scores. The fMRI contrast beta weights are standardized values, and EXT and CTQ scores were converted to Z scores. The binary outcome was the presence vs. absence of a past-year DSM-IV diagnosis from age 14, 16, or 19 years old. Mediation and moderation analyses were performed using the PROCESS macro for SPSS by Andrew F. Hayes (41).

The Box-Tidwell test was used to assess the linearity of each continuous variable with the logit of the dependent variable (42). A Bonferroni correction was applied with all terms in the model, resulting in an adjusted alpha level of *p* < 0.0083 (43). All continuous variables were linearly related to the logit of the dependent variable.

Receiver operating characteristic (ROC) curves were plotted for each model. The sensitivity (true positive rate) is plotted on the y-axis and 1-specificity (false positive rate) is plotted on the x-axis. The area under the curve (AUC) assesses the strength of the overall model, where an AUC near 1 reflects a good model and an AUC near 0.5 reflects poor model discrimination (44).

## RESULTS

### Demographic Characteristics

The participants were 637 males (47.6%) and 701 females (52.4%). Among these 1338 participants, 29.4% were diagnosed with one or more DSM-IV psychiatric disorders at ages 14, 16, or 19 years of age. The most common diagnoses were mood and anxiety disorders, followed by eating disorders, attention deficit-hyperactivity disorder (ADHD), and conduct disorder (Table 1). At the age-14 interview, 12.9% (147/1135, 3 missing data) of participants had one or more confirmed disorders; at age 16, 16.9% (204/1209, 129 missing data) had one or more disorders; and at age 19, 16.2% (212/1308, 30 missing data) had one or more disorders. Among the 1183 individuals who completed the DAWBA at all three visits, 230 (19.4%) were diagnosed with a disorder at a single timepoint, 91 (7.7%) were diagnosed at two timepoints, and 29 (2.5%) received diagnoses at all three timepoints.

**Table 1.** Participant demographic characteristics.

|  |  | **n (%)** |
| --- | --- | --- |
| **Sex** | Male | 637 (47.6) |
|  | Female | 701 (52.4) |
| **Language** | German | 591 (44.2%) |
|  | English | 568 (42.4) |
|  | French | 179 (13.4%) |
| **DAWBA Diagnoses** | Anxiety | 191 (14.3%) |
|  | Depression | 157 (11.7%) |
|  | Eating disorder | 57 (4.2%) |
|  | ADHD | 51 (3.8%) |
|  | Conduct disorder | 48 (3.6%) |
|  | PTSD | 23 (1.7%) |
|  | Tic disorder | 17 (1.3%) |
|  | OCD | 16 (1.2%) |
|  | Autism | 9 (0.7%) |
|  | Psychosis | 3 (0.2%) |
|  | Other | 11 (0.8%) |
| **Score (mean; SD)** | CTQ | 31.89 (7.98) |
|  | EXT | 3.33 (1.05) |

### Age-14 models

All tested binary regression models were statistically significant in predicting who had a psychiatric disorder by age 19 (*p* < 1.0 x 10^-21^). EXT and CTQ contributed significantly to all models (*p* < 0.001; see eTables 1-7 in Supplement). In models with the ‘anticipated large – no win’ contrast, the caudate and VS were significant individual predictors (Table 2). For models that included the ‘large – small win’ contrast, the caudate, VS, putamen, and ACC emerged as predictors (Table 2). These models explained over 12.6% of the variance in DSM-IV disorders (Nagelkerke *R*^2^). Sensitivity and specificity of the models ranged from 17.0-19.7% and 95.8- 96.7%, respectively, with ROC AUC ranging from 0.68 to 0.69. In each model, a DSM-IV diagnosis by age 19 was predicted by higher EXT and CTQ scores and lower mesocorticolimbic responses. Sensitivity of the three-factor models was similar when the outcome included participants with a diagnosis at any one timepoint, compared to individuals with diagnoses at multiple timepoints (see eResults Supplement).

**Table 2.** Binomial regression models with each ROI and contrast.

| **Contrast** | **Reward Anticipation**<br>**fMRI response**<br>**(p-value, OR, 95% CI)** | **EXT**<br>**(p-value, OR,**<br>**95% CI)** | **CTQ**<br>**(p-value, OR,**<br>**95% CI)** | **Full model**<br>**(p-value,**<br>**Nagelkerke**<br>**R<sup>2</sup>)** |
| --- | --- | --- | --- | --- |
| **Large –**<br>**No win** | Age 14 L Caudate:<br>0.019, 0.775<br>(0.626-0.959) | 1.64 x 10<sup>-8</sup>, 1.481<br>(1.292-1.697) | 1.07 x 10<sup>-11</sup>, 1.673<br>(1.442-1.940) | 7.32 x 10<sup>-24</sup>,<br>0.132 |
|  | Age 14 R Caudate:<br>0.013, 0.773<br>(0.632-0.946) | 1.30 x 10<sup>-8</sup>, 1.486<br>(1.296-1.703) | 9.87 x 10<sup>-12</sup>, 1.671<br>(1.441-1.937) | 5.13 x 10<sup>-24</sup>,<br>0.133 |
|  | Age 14 L VS:<br>0.017, 0.699<br>(0.521-0.939) | 1.03 x 10<sup>-8</sup>, 1.489<br>(1.299-1.706) | 5.97 x 10<sup>-12</sup>, 1.683<br>(1.451-1.953) | 6.82 x 10<sup>-24</sup>,<br>0.133 |
|  | Age 19 R VS:<br>0.025, 0.734<br>(0.559-0.962) | 1.26 x 10<sup>-7</sup>, 1.440<br>(1.258-1.649) | 8.63 x 10<sup>-12</sup>, 1.681<br>(1.448-1.951) | 6.94 x 10<sup>-22</sup>,<br>0.118 |
| **Large –**<br>**Small**<br>**win** | Age 14 R Caudate:<br>0.050, 0.776<br>(0.603-0.999) | 1.10 x 10<sup>-8</sup>, 1.481<br>(1.294-1.694) | 6.41 x 10<sup>-12</sup>, 1.666<br>(1.440-1.928) | 1.73 x 10<sup>-23</sup>,<br>0.127 |
|  | Age 14 R VS:<br>0.019, 0.633<br>(0.432-0.928) | 4.68 x 10<sup>-9</sup>, 1.496<br>(1.307-1.712) | 3.80 x 10<sup>-12</sup>, 1.674<br>(1.447-1.936) | 6.56 x 10<sup>-24</sup>,<br>0.129 |
|  | Age 14 L Putamen:<br>0.043, 0.747<br>(0.563-0.991) | 8.69 x 10<sup>-9</sup>, 1.484<br>(1.297-1.698) | 3.72 x 10<sup>-12</sup>, 1.673<br>(1.447-1.934) | 1.53 x 10<sup>-23</sup>,<br>0.127 |
|  | Age 14 ACC:<br>0.005, 0.710<br>(0.558-0.904) | 5.93 x 10<sup>-9</sup>, 1.493<br>(1.305-1.709) | 4.63 x 10<sup>-12</sup>, 1.672<br>(1.445-1.934) | 2.40 x 10<sup>-24</sup>,<br>0.131 |

### Age-19 models

Each binary regression model was significant, and EXT and CTQ contributed to all models (*p* < 0.001) (see eTable 8 in Supplement). For the model that included the ‘large – no win’ contrast, the VS was also an individual predictor (*p* = 0.025; Table 2) and the overall model had a predictive accuracy of 73.5% (χ ^2^(3) = 101.63, *p* = 6.94 x 10^-22^). The model explained 11.8% of the variance in diagnoses (Nagelkerke *R*^2^). Sensitivity and specificity of the model were 17.7% and 96.7%, respectively, and it had an ROC AUC of 0.67. As above, DSM-IV diagnoses were predicted by higher EXT and CTQ scores and lower fMRI responses. There were no significant predictors for the ‘large – small win’ or ‘small – no win’ conditions.

### Effect of Sex

For each model where the fMRI contrast was a significant individual predictor, sex was added as a fourth factor (see eTables 9-16 in Supplement). Sex was a significant factor in every model (*p* < 1.0 x 10^-6^), where being female doubled the odds of being diagnosed with a disorder (e.g., for the age 14 left VS model, OR = 2.202, 95% CI 1.661-2.919). Its addition also increased each model’s statistical significance (*p* < 1.0 x 10^-26^), now explaining >14% of the variance in DSM-IV diagnoses (Nagelkerke *R^2^*).

### Mediation and Moderation Analyses

To investigate whether the three factors influenced each other, we tested an interaction term for CTQ scores at low (-1 SD), mean, and high (+1 SD) levels of the fMRI repsonse. At age 14, ACC (*p* = 0.0038) and left putamen (*p* = 0.0135) interacted with CTQ scores; the lower the reward anticipation response, the greater the effect of high CTQ scores on the probability of a disorder by age 19 (Figure 1).

Mediation analyses found that the association between high CTQ scores and the probability of a disorder was carried by two pathways, a direct pathway (*B* = 0.477, 95% confidence interval = 0.345 to 0.609) and an indirect pathway whereby the CTQ-disorder association was mediated by EXT scores (*a* x *b* indirect effect: *B* = 0.0535, 95% confidence interval = 0.0301 to 0.0835; Figure 2).

**Figure 2.**
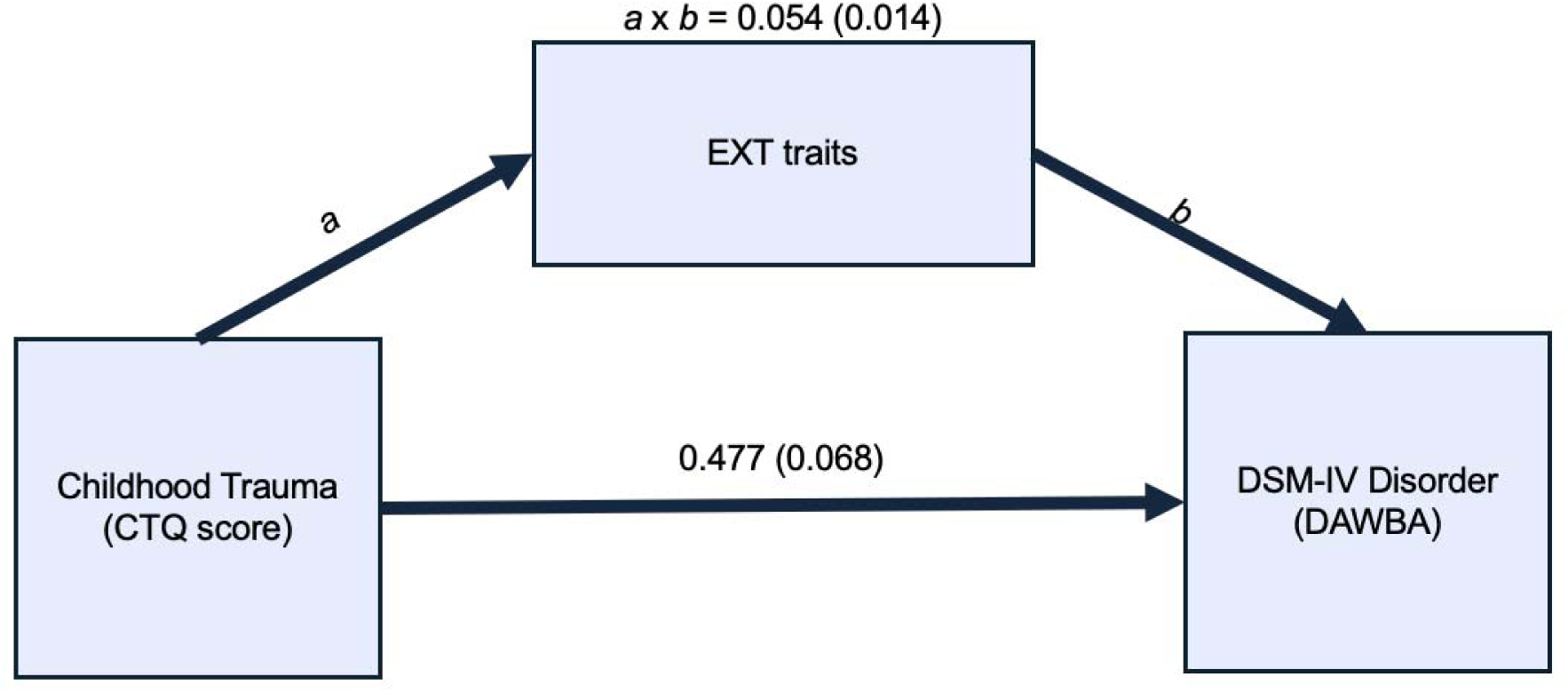
Mediation of Childhood Trauma by Externalizing Traits Mediation analysis results. *a* x *b* shows the indirect effects (SE) of CTQ scores on the odds of DAWBA identified DSM-IV disorders via higher EXT. Direct effects of CTQ scores on the odds of a DSM-IV disorder (ß (SE)) are given with the direct arrow.

### Substance Use Disorders

Substance use disorder (SUD) diagnoses derived from the Structured Clinical Interview for the DSM-IV (SCID; 45) SUD screening module were added to the DAWBA diagnoses, and each model was run again (see eMethods in Supplement). At age 14, smaller fMRI responses for the ‘large – no win’ contrast in the left VS (*p* = 0.050) and right caudate (*p* = 0.031) were significant individual predictors of any diagnosis (SUD + DAWBA) by age 19. Similarly, psychopathology was predicted by smaller age 14 ‘large – small win’ contrast responses in the ACC (*p* = 0.045), with other regions nearing significance (see eTables 17-23 in Supplement).

There were no significant predictors for the ‘small – no win’ contrast.

With age 19 fMRI data, smaller responses in the right VS (*p* = 0.007) and right caudate (*p* = 0.014) were significant individual predictors of psychopathology for the ‘large – no win’ contrast. For the ‘small – no win’ contrast, smaller responses in the left caudate (*p* = 0.022) were a significant predictor (see eTables 24-26 in Supplement). There were no significant predictors for the ‘large – small win’ contrast.

Sex remained significant when added as a fourth factor. At age 19, fMRI effects were seen throughout the striatum. At age 14, fMRI effects were seen in the midbrain, striatum, and ACC (see eTables 27-36 in Supplement).

## DISCUSSION

The study’s primary objective was to test our recently proposed diathesis-stress model (27) in a large population-based cohort. As hypothesized, we found that a combination of greater adolescent EXT behavior, more childhood trauma, and decreased mesocorticolimbic reward-anticipation responses, as early as age 14, identified young adults with a history of psychopathology. Together, these findings delineate specific features of a novel diathesis-stress model of commonly comorbid early onset disorders.

The statistical effect of each individual factor was modest and associated with roughly 30 to 60% changes in odds ratios for a psychiatric disorder. Together, though, the full model was highly significant, and this was seen when incorporating activation responses throughout mesocorticolimbic sites (*p* < 10^-21^).

The associations with early life adversity were moderated by responses in the striatum and anterior cingulate cortex (ACC). Low reward anticipation responses at age 14 were associated with a stronger relationship between CTQ scores and the likelihood of a psychiatric disorder by age 19. Large reward anticipation responses, in comparison, weakened the association between early life adversity and psychiatric disorders, plausibly providing protection. One intriguing possibility is that this contribution of low striatal reward anticipation reflects a transdiagnostic association with anhedonia (46; Supplement). The ACC, in comparison, has been implicated as an interface between motor control, cognition and motivational drive, the latter mediated by subcortical monoaminergic systems (47).

Consistent with previous evidence (48, 49), early life adversity increased the likelihood of a psychiatric disorder through two pathways, a direct pathway and an indirect pathway mediated by higher EXT behavior. Higher EXT behavior during adolescence was associated with a higher risk for both EXT (e.g., SUDs, ADHD) and internalizing disorders (e.g., mood and anxiety disorders). This too is consistent with our previous study (27) and the larger literature (50, 1, 3).

In our original three-factor model, psychiatric disorders were predicted by lower levels of inhibitory dopamine autoreceptor availability, as assessed by PET. Our present findings of weaker fMRI responses to reward anticipation might seem contradictory, but both higher and lower fMRI responses have been reported in people with psychiatric disorders, compared with healthy controls (51). The direction of these associations may be related to the type of challenge presented (52). In the presence of monetary reward cues, psychiatric disorders are predicted by weaker mesocorticolimbic responses (22, 53, 26, 54). In the presence of more disease-specific cues, larger fMRI responses are seen in high-risk participants (55, 56, 57). Very recently, we tested this hypothesis in the context of our diathesis-stress model. In youth followed since birth (n = 44), the combination of higher adolescent EXT scores, higher early life adversity, and *stronger* mesocorticolimbic activations to *alcohol cues* at age 18 predicted more alcohol use problems at age 25 (58). Together, these results suggest that the three-factor model incorporating responses to disease-independent cues predicts a wide range of commonly comorbid early onset disorders. In comparison, responses to disease-related cues can predict susceptibility to more specific outcomes. Both effects might be seen most clearly after controlling for the influence of early life adversity (7, 59) and adolescent behavioral tendencies (50, 1, 3). Together, these observations identify components of a hypothesized hierarchical model of psychopathology whereby general factors increase susceptibility to commonly comorbid disorders and superimposed features increase risk for progressively more narrow symptoms and problems (60, 61; Figure 3). Indeed, the biased attachment of mesocorticolimbic activations to a narrow set of cues might shape many pathological behaviors, accounting for specific symptoms and disorders (51).

**Figure 3.**
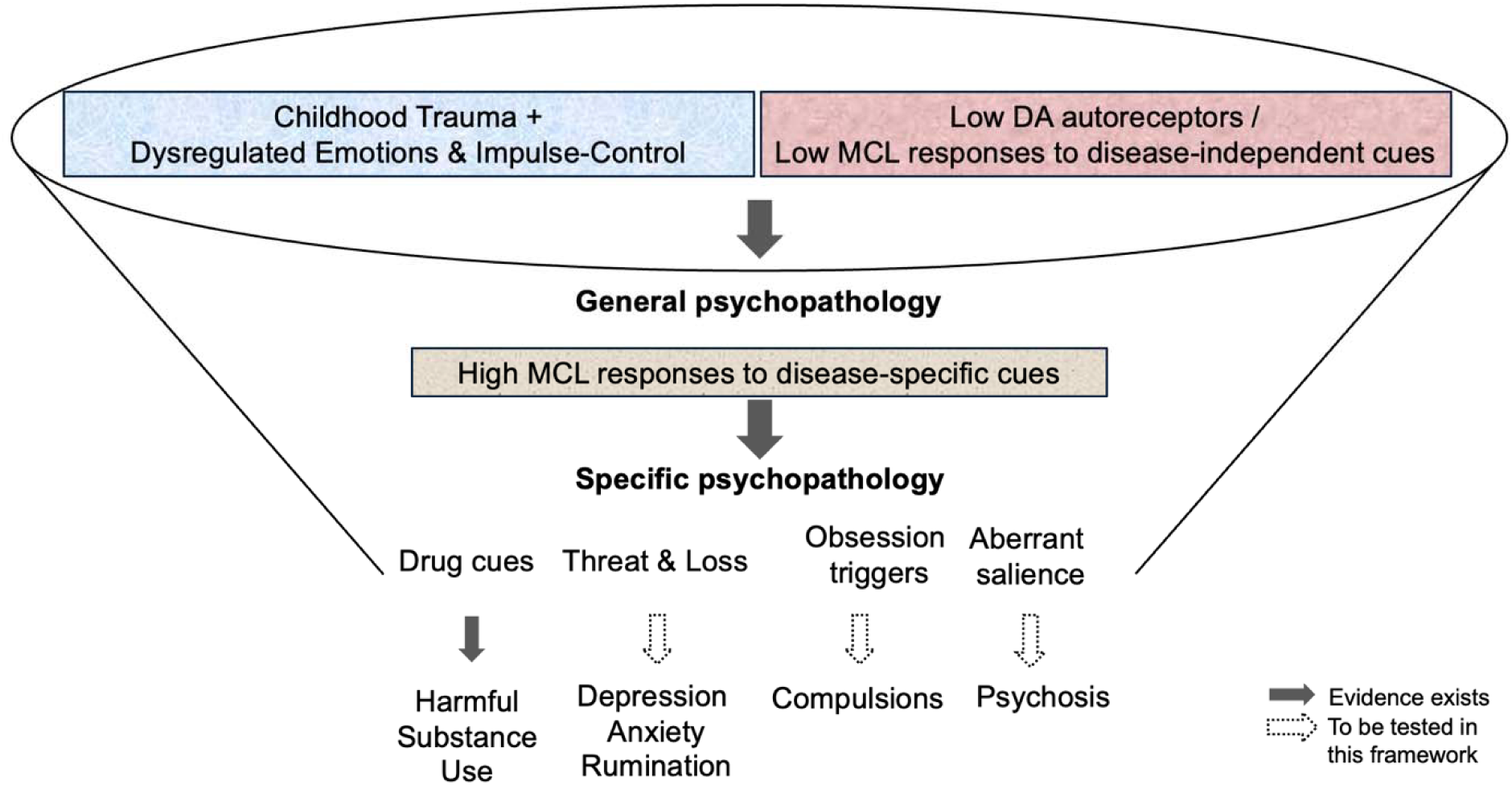
A Hierarchical Diathesis-Stress Model The model proposes that a broad liability to diverse psychopathology is promoted by a combination of (i) early life trauma, (ii) dysregulated emotional and impulse control, and (iii) poorly regulated mesocorticolimbic reactivity as indexed by low dopamine (DA) autoreceptor availability and low mesocorticolimbic (MCL) responses to disease-independent cues. Specific disorders are predicted by high mesocorticolimbic responses to disease-specific cues after controlling for the first two factors. The solid arrows identify pathways with empirical support.

## Strengths and Limitations

The present study tested a novel diathesis-stress model. It benefitted from having a large sample size and longitudinally collected behavioral, diagnostic, and neuroimaging data throughout adolescence. The cohort was also large enough to test statistical interactions between the predictors. These strengths noted, the following features should be considered. First, sensitivity in the current study was lower than that seen in our PET-based model (17-19.7% vs 75%, respectively). This observation raises the possibility that the neurotransmitter specific information provided by PET (compared with fMRI) might better identify the relevant disease-related processes. Alternatively, some of the cases classified here as “false negatives” might reflect a tendency of the DAWBA to over-diagnose (32) whereas those identified as “false positives” might have experienced mental health problems either not captured by the DAWBA interview (see eMethods in Supplement) or outside of the DAWBA’s temporal window (ages 13 – 19 y.o.); in the PET study, the full range of DSM-5 diagnoses were available from birth to 18 y.o. Each of these influences would decrease the chances of identifying an effect in the present study, raising the possibility that the true effects are larger than reported. Despite these potential issues, the DAWBA is considered well-validated for large sample studies (32), sensitivity increased when sex was added as a fourth factor, and both prediction accuracy and specificity seen here were high and similar to what was found in the PET study. Second, the CTQ was administered at age 19 and might have been affected by recall bias. While some reports have advised caution with the physical neglect subscale (e.g., 62), the CTQ has consistently demonstrated good reliability (62, 63), convergent validity with other measures of childhood maltreatment (64), and consistency across adolescence (65). Still, there is evidence that the association between reported childhood trauma and psychopathology is influenced by subjective experiences (66, 67), potentially further revealing individual differences in the impact of early life adversity. These effects noted, there is compelling evidence that early life adversity has effects on corticostriatal reactivity (59) and psychiatric outcomes (68, 7) that are independent of subjective recall. Third, the present study identified an effect of sex. While there are differences in the prevalence rates of specific disorders across sex, the lifetime prevalence of psychiatric problems is similar across males and females (69). Despite this, adding sex as a fourth factor strengthened our model, with female sex increasing the likelihood of having a disorder by age 19 even in models that included SUD diagnoses. This might reflect differences in the age of onset of different disorders (70) or be due to the most prevalent disorders in our sample (depression, anxiety, eating disorders) being more common in females (71). Fourth, repeated administrations of the MID task might habituate participants to the parameters (72, 26). Modifying reward magnitude across waves has been proposed as a solution to changes in motivation over time (72).

## Conclusion

Further work will be needed to establish causality, but this novel diathesis-stress model identifies transdiagnostic predictors of psychopathology – the present study suggests that these predictions benefit from the inclusion of multi-level, biopsychosocial factors.

## Acknowledgements

### Author Contributions

Caswell, Hosseini-Kamkar and Leyton had full access to all the data in the study and takes responsibility for the integrity of the data and the accuracy of the data analysis.

Concept and design: Leyton

*Acquisition, analysis, or interpretation of the data:* Caswell, Hosseini-Kamkar, Cox, Palacio Prada, Iqbal, Nikolic, Banaschewski, Barker, Bokde, Brühl, Desrivières, Flor, Garavan, Gowland, Grigis, Heinz, Martinot, Paillère Martinot, Artiges, Nees, Papadopoulos Orfanos, Poustka, Smolka, Hohmann, Vaidya, Walter, Whelan, Schumann, Paus, Leyton

Drafting of the manuscript: Caswell, Leyton

*Critical revision of the manuscript for important intellectual content:* Hosseini-Kamkar, Cox, Palacio Prada, Iqbal, Nikolic, Banaschewski, Barker, Bokde, Brühl, Desrivières, Flor, Garavan, Gowland, Grigis, Heinz, Martinot, Paillère Martinot, Artiges, Nees, Papadopoulos Orfanos, Poustka, Smolka, Hohmann, Vaidya, Walter, Whelan, Schumann, Paus, Leyton

*Statistical analysis:* Caswell, Hosseini-Kamkar, Cox, Leyton

*Obtained funding:* Bokde, Desrivières, Flor, Heinz, Martinot, Paillère Martinot, Whelan, Schumann, Paus, Leyton

*Administrative, technical, or material support:* Bokde, Desrivières, Flor, Grigis, Gowland, Heinz, Brühl, Martinot, Nees, Papadopoulos Orfanos, Smolka, Vaidya, Walter, Whelan, Schumann

*Supervision:* Hosseini-Kamkar, Cox, Paus, Leyton

### Conflict of interest disclosures

Dr Banaschewski served in an advisory or consultancy role for AGB Pharma, eye level, Infectopharm, Medice, Neurim Pharmaceuticals, Oberberg GmbH and Takeda. He received conference support or speaker’s fee by Janssen-Cilag, Medice and Takeda. He received royalities from Hogrefe, Kohlhammer, CIP Medien, Oxford University Press; the present work is unrelated to these relationships. Dr Barker has received honoraria from General Electric Healthcare for teaching on scanner programming courses. Dr Poustka served in an advisory or consultancy role for Roche and Viforpharm and received a speaker’s fee by Shire. She received royalties from Hogrefe, Kohlhammer and Schattauer.

### Funding/Support

This work received support from the following sources: the European Union-funded FP6 Integrated Project IMAGEN (Reinforcement-related behaviour in normal brain function and psychopathology) (LSHM-CT- 2007-037286), the Horizon 2020 funded ERC Advanced Grant ‘STRATIFY’ (Brain network based stratification of reinforcement-related disorders) (695313), Horizon Europe ‘environMENTAL’, grant no: 101057429, UK Research and Innovation (UKRI) Horizon Europe funding guarantee (10041392 and 10038599), Human Brain Project (HBP SGA 2, 785907, and HBP SGA 3, 945539), the Chinese government via the Ministry of Science and Technology (MOST), The German Center for Mental Health (DZPG), the Bundesministerium für Bildung und Forschung (BMBF grants 01GS08152; 01EV0711; Forschungsnetz AERIAL 01EE1406A, 01EE1406B; Forschungsnetz IMAC-Mind 01GL1745B), the Deutsche Forschungsgemeinschaft (DFG project numbers 458317126 [COPE], 186318919 [FOR 1617], 178833530 [SFB 940], 386691645 [NE 1383/14-1], 402170461 [TRR 265], 454245598 [IRTG 2773]), the Medical Research Foundation and Medical Research Council (grants MR/R00465X/1 and MR/S020306/1), the National Institutes of Health (NIH) funded ENIGMA-grants 5U54EB020403-05, 1R56AG058854-01 and U54 EB020403 as well as NIH R01DA049238, the National Institutes of Health, Science Foundation Ireland (16/ERCD/3797). NSFC grant 82150710554. Further support was provided by grants from the Agence Nationale de la Recherche (ANR-12-SAMA-0004, AAPG2019 - GeBra), the Eranet Neuron (AF12- NEUR0008-01 - WM2NA; and ANR-18-NEUR00002-01 - ADORe), the Fondation de France (00081242), the Fondation pour la Recherche Médicale (DPA20140629802), the Mission Interministérielle de Lutte-contre-les-Drogues-et-les-Conduites-Addictives (MILDECA), the Assistance-Publique-Hôpitaux-de-Paris and INSERM (interface grant), Paris Sud University IDEX 2012, the Fondation de l’Avenir (grant AP-RM-17-013), the Fédération pour la Recherche sur le Cerveau. ImagenPathways “Understanding the Interplay between Cultural, Biological and Subjective Factors in Drug Use Pathways” is a collaborative project supported by the European Research Area Network on Illicit Drugs (ERANID). This study is based on independent research commissioned and funded in England by the National Institute for Health Research (NIHR) Policy Research Programme (project ref. PR-ST-0416-10001). Dr. Leyton was supported by the Canadian Institutes for Health Research (CIHR; Grant Number MOP-453327). Caswell was supported by a scholarship from CIHR.

### Role of the Funder/Sponsor

The funding sources had no role in the design and conduct of the study; collection, management, analysis, and interpretation of the data; preparation, review, or approval of the manuscript; and decision to submit the manuscript for publication.

### IMAGEN consortium members

Tobias Banaschewski, MD, PhD, Gareth J. Barker, PhD, Arun L.W. Bokde, PhD, Rüdiger Brühl, PhD, Sylvane Desrivières, PhD, Herta Flor, PhD, Hugh Garavan, PhD, Penny Gowland, PhD, Antoine Grigis, PhD, Andreas Heinz, MD, PhD, Jean-Luc Martinot, MD, PhD, Maire-Laure Paillère Martinot, MD, Eric Artiges, MD, PhD, Frauke Nees, MD, PhD, Dimitri Papadopoulos Orfanos, PhD, Luise Poustka, MD, Michael N. Smolka, MD, Sarah Hohmann, PhD, Nilakshi Vaidya, MSc, Henrik Walter, MD, PhD, Robert Whelan, PhD, Gunter Schumann, MD, Tomáš Paus, MD, PhD

### Disclaimer

The views expressed in this article are those of the authors and not necessarily those of the national funding agencies or ERANID.

## Data Availability

All data in the present work are available upon request to the IMAGEN consortium.

https://www.imagen-project.org/dataaccess

## Notes

### Author Declarations

The institutional ethics committe of King's College London, University of Nottingham, Trinity College Dublin, Technische Universitat Dresden, Commissariat a l'Energie Atomique et aux Energies Alternatives, University Medical Center at the University of Hamburg and the University of Heidelberg gave ethical approval for this work.

