## Supplement for "A Novel Diathesis-Stress Model of Comorbid Early Onset Psychiatric Disorders"

**eMethods**

*Diagnoses*

DSM-IV psychiatric disorders were assessed at 14, 16, and 19 years of age using the Development and Wellbeing Assessment (DAWBA; 1). The DAWBA uses six probability bands ranging from <0.1% likelihood to >70% likelihood of having a given disorder in the past year. These computer-generated predictions were reviewed by clinicians who either accepted or rejected the computed diagnosis. Diagnoses assessed by the DAWBA include separation anxiety, specific phobia, social phobia, panic disorder, agoraphobia, post-traumatic stress disorder, obsessive compulsive disorder, generalized anxiety disorder, body dysmorphic disorder, disruptive mood dysregulation disorder, major depression, attention deficit-hyperactivity disorder/hyperkinesis, oppositional defiant disorder, conduct disorder, eating disorders (anorexia, bulimia, binge eating disorder), autism spectrum disorders, tic disorders, and bipolar disorders. Clinician-reviewed scores for each time point were used, and participants were coded as either ‘1’ (one or more disorders at one or more time points) or ‘0’ (no disorder at any time point).

The DAWBA does not assess for substance use disorders (SUDs). Instead, at ages 16 and 19, participants completed the Structured Clinical Interview for the DSM-IV (SCID; 2) SUD screening module. Individuals who met criteria for substance abuse or dependence on the SCID-SUD screening at age 16 or 19 were coded as ‘1’. Individuals who did not meet SUD criteria at any time point were coded as ‘0’. These constructed SUD diagnoses were added to the clinician-rated DAWBA diagnoses, so that an individual with a DAWBA DSM-IV disorder and/or SUD is coded as 1, and an individual with neither at any time point is coded as 0. When SUDs were included, 453 participants met criteria for a DAWBA DSM-IV disorder, SUD, or both by age 19. Only 59 participants met criteria for a SUD alone.

*Externalizing traits*

The items from the hyperactivity and conduct subscales of the Strengths and Difficulty questionnaire were selected to create a harmonized version of our original externalizing measure (EXT; 3), originally developed from the Social Behaviour Questionnaire (SBQ; 4).

*Monetary Incentive Delay Task*

Prior to the imaging session, participants learned how to perform the monetary incentive delay (MID; 5) task outside the scanner. The task begins by presenting an incentive cue on the left or right side of the screen for 250 ms. The cue indicates the points to be earned in a given trial. The three possible reward magnitudes are 0 points (no reward condition), 2 points (small reward condition), and 10 points (large reward condition). After the cue presentation, the screen is blank for 4-4.5 s. The target screen is then presented, and participants must respond to the target by pressing a button in their left or right hand. The timing of the target presentation is altered to achieve a 66% success rate for each participant. After target presentation the screen displays the points won in the current trial and the accumulated points from previous trials. During the intertrial interval of 3.5-4.15 s, participants focus on a fixation cross. There are 66 trials in total, 22 per reward condition.

**Regions of Interest**

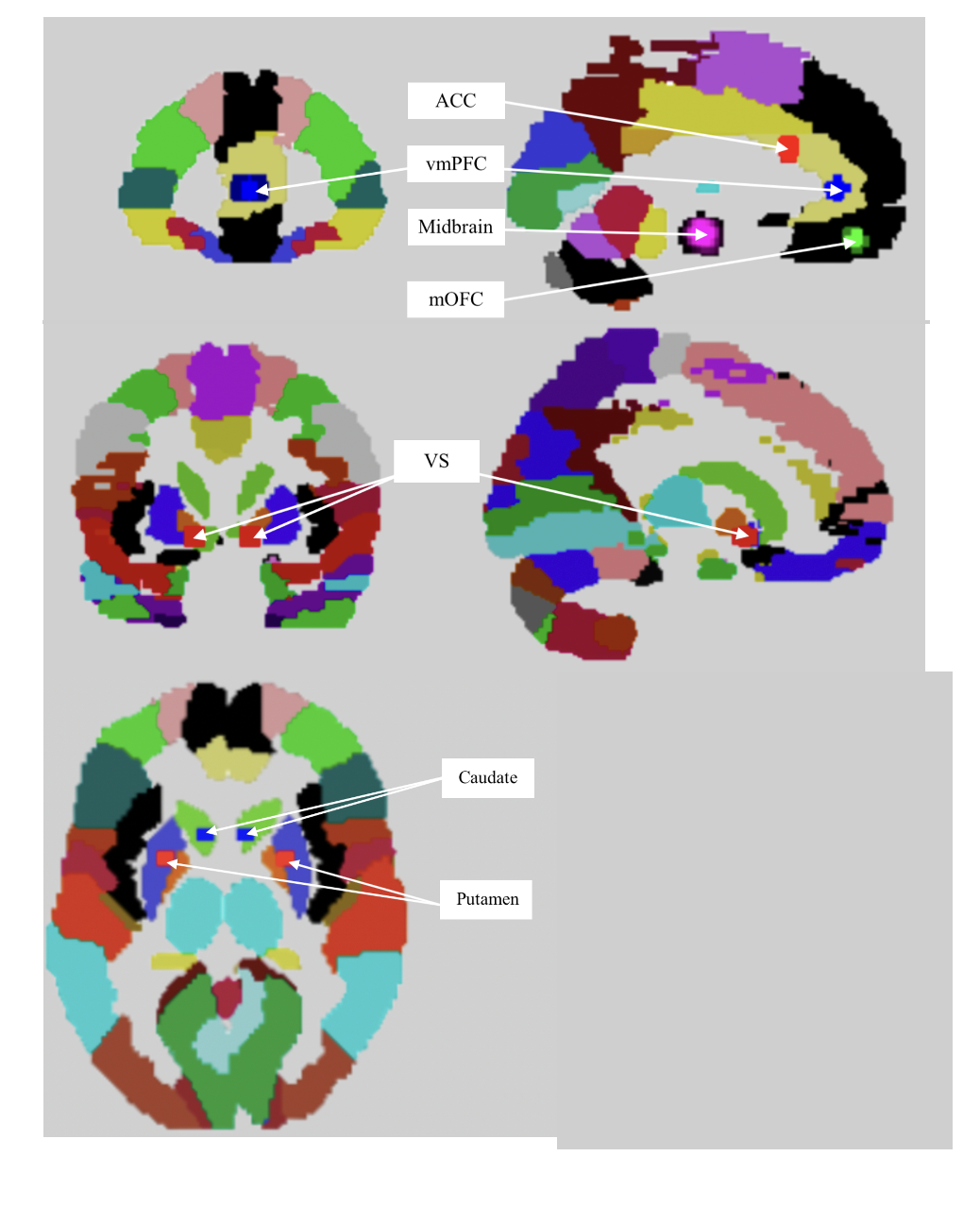

**eFigure 1**. Region of interest masks (x, y, z). Anterior cingulate cortex (ACC; 0, 20, 25); ventromedial prefrontal cortex (vmPFC; Left: -3, 41, 7; Right: 3, 41, 7); midbrain (Duke Atlas; 6); medial orbitofrontal cortex (mOFC; Left: -2. 5-, -16; Right: 2, 48, -14); ventral striatum (VS; Left: -12, 10, -6; Right: 12, 10, -6); caudate (Left: -8, 14, 2; Right: 8, 14, 2); putamen (Left: -24, 4, 6; Right: 24, 4, 0).

**eResults**

*Diagnoses at multiple timepoints*

To determine whether the model sensitivity differed when predicting individuals with a disorder at more than one age, we re-ran the three-factor models, now with diagnoses restricted to individuals with any disorder at one or more timepoints. The sensitivity in these models was similar to the original three-factor models, with a range from 17.2-20.1% compared with 17-19.7% in the original models. This suggests that the model has comparable strength in accurately identifying individuals with a disorder at one timepoint or individuals with a disorder at multiple timepoints.

*Complete model information for the three-factor models predicting one or more DSM-IV disorders from ages 14-19*

**eTable 1.** Complete three-factor model results including the age-14 BOLD response to the anticipated ‘Large – no win’ condition in the left ventral striatum (VS).

| **N = 1145** | | **Left VS BOLD**  **Age 14 Large – No win**  **(-2.3 to 2.6)** | **EXT score**  **(-2.7 to 3.9)** | **CTQ**  **(-0.9 to 8.2)** |
| --- | --- | --- | --- | --- |
| **Beta ± S.E.** | | -0.358 (0.150) | 0.398 (0.07) | 0.521 (0.08) |
| **Wald** | | 5.667 | 32.780 | 47.340 |
| **p** | | 0.017 | 1.03 x10^-8^ | 5.97 x10^-12^ |
| **Odds Ratio** | | 0.699 | 1.489 | 1.683 |
| **95% CI for Odds Ratio** | **Lower** | 0.521 | 1.299 | 1.451 |
|  | **Upper** | 0.939 | 1.706 | 1.953 |
| **Chi squared** | | | χ^2^(3) = 110.960, p = 6.82 x10^-24^ | |
| **Nagelkerke R2 & AUC** | | | 0.133 & 68.7% | |
| **Classification predictive accuracy** | | | 74.6% | |
| **Sensitivity & Specificity** | | | 19.7% & 96.3% | |
| **Positive & Negative Predictive value** | | | 68.1% & 75.2% | |

**eTable 2.** Complete three-factor model results including the age-14 BOLD response to the anticipated ‘Large – no win’ condition in the left caudate.

| **N = 1145** | | **Left caudate BOLD**  **Age 14 Large – No win**  **(-2.4 to 4.4)** | **EXT score**  **(-2.7 to 3.9)** | **CTQ**  **(-0.9 to 8.2)** |
| --- | --- | --- | --- | --- |
| Beta ± S.E. | | -0.255 (0.109) | 0.393 (0.07) | 0.514 (0.08) |
| Wald | | 5.518 | 31.881 | 46.205 |
| p | | 0.019 | 1.64 x10^-8^ | 1.07 x10^-11^ |
| Odds Ratio | | 0.775 | 1.481 | 1.673 |
| **95% CI for Odds Ratio** | Lower | 0.626 | 1.292 | 1.442 |
|  | Upper | 0.959 | 1.697 | 1.940 |
| **Chi squared** | | | χ^2^(3) = 110.818, p = 7.32 x10^-24^ | |
| **Nagelkerke R2 & AUC** | | | 0.132 & 68.7% | |
| **Classification predictive accuracy** | | | 74.2% | |
| **Sensitivity & Specificity** | | | 18.5% & 96.3% | |
| **Positive & Negative Predictive value** | | | 66.7% & 74.9% | |

**eTable 3.** Complete three-factor model results including the age-14 BOLD response to the anticipated ‘Large – no win’ condition in the right caudate.

| **N = 1145** | | **Right caudate BOLD**  **Age 14 Large – No win**  **(-2.2 to 3.1)** | **EXT score**  **(-2.7 to 3.9)** | **CTQ**  **(-0.9 to 8.2)** |
| --- | --- | --- | --- | --- |
| Beta ± S.E. | | -0.257 (0.103) | 0.396 (0.07) | 0.513 (0.08) |
| Wald | | 6.238 | 32.338 | 46.354 |
| p | | 0.013 | 1.30 x10^-8^ | 9.87 x10^-12^ |
| Odds Ratio | | 0.773 | 1.486 | 1.671 |
| **95% CI for Odds Ratio** | Lower | 0.632 | 1.296 | 1.441 |
|  | Upper | 0.946 | 1.703 | 1.937 |
| **Chi squared** | | | χ^2^(3) = 111.534, p = 5.13 x10^-24^ | |
| **Nagelkerke R2 & AUC** | | | 0.133 & 68.5% | |
| **Classification predictive accuracy** | | | 74.7% | |
| **Sensitivity & Specificity** | | | 19.7% & 96.5% | |
| **Positive & Negative Predictive value** | | | 68.8% & 75.2% | |

**eTable 4.** Complete three-factor model results including the age-14 BOLD response to the anticipated ‘Large – small win’ condition in the anterior cingulate cortex (ACC).

| **N = 1178** | | **ACC BOLD**  **Age 14 Large – Small win**  **(-3.6 to 3.5)** | **EXT score**  **(-2.7 to 3.9)** | **CTQ**  **(-0.9 to 8.2)** |
| --- | --- | --- | --- | --- |
| Beta ± S.E. | | -0.342 (0.123) | 0.401 (0.07) | 0.514 (0.07) |
| Wald | | 7.741 | 33.858 | 47.839 |
| p | | 0.005 | 5.93 x10^-9^ | 4.63 x10^-12^ |
| Odds Ratio | | 0.710 | 1.493 | 1.672 |
| **95% CI for Odds Ratio** | Lower | 0.558 | 1.305 | 1.445 |
|  | Upper | 0.904 | 1.709 | 1.934 |
| **Chi squared** | | | χ^2^(3) = 113.071, p = 2.40 x10^-24^ | |
| **Nagelkerke R2 & AUC** | | | 0.131 & 68.4% | |
| **Classification predictive accuracy** | | | 73.4% | |
| **Sensitivity & Specificity** | | | 17.0% & 95.8% | |
| **Positive & Negative Predictive value** | | | 62.0% & 74.4% | |

**eTable 5.** Complete three-factor model results including the age-14 BOLD response to the anticipated ‘Large – small win’ condition in the left putamen.

| **N = 1178** | | **Left putamen BOLD**  **Age 14 Large – Small win**  **(-2.0 to 3.9)** | **EXT score**  **(-2.7 to 3.9)** | **CTQ**  **(-0.9 to 8.2)** |
| --- | --- | --- | --- | --- |
| Beta ± S.E. | | -0.292 (0.144) | 0.395 (0.07) | 0.514 (0.07) |
| Wald | | 4.089 | 33.115 | 48.271 |
| p | | 0.043 | 8.69 x10^-9^ | 3.72 x10^-12^ |
| Odds Ratio | | 0.747 | 1.484 | 1.673 |
| **95% CI for Odds Ratio** | Lower | 0.563 | 1.297 | 1.447 |
|  | Upper | 0.991 | 1.698 | 1.934 |
| **Chi squared** | | | χ^2^(3) = 109.333, p = 1.53 x10^-23^ | |
| **Nagelkerke R2 & AUC** | | | 0.127 & 68.1% | |
| **Classification predictive accuracy** | | | 74.5% | |
| **Sensitivity & Specificity** | | | 18.8% & 96.7% | |
| **Positive & Negative Predictive value** | | | 69.2% & 75.0% | |

**eTable 6.** Complete three-factor model results including the age-14 BOLD response to the anticipated ‘Large – small win’ condition in the right ventral striatum (VS).

| **N = 1178** | | **Right VS BOLD**  **Age 14 Large – Small win**  **(-1.7 to 2.2)** | **EXT score**  **(-2.7 to 3.9)** | **CTQ**  **(-0.9 to 8.2)** |
| --- | --- | --- | --- | --- |
| Beta ± S.E. | | -0.458 (0.195) | 0.403 (0.07) | 0.515 (0.07) |
| Wald | | 5.502 | 34.317 | 48.227 |
| p | | 0.019 | 4.68 x10^-9^ | 3.80 x10^-12^ |
| Odds Ratio | | 0.633 | 1.496 | 1.674 |
| **95% CI for Odds Ratio** | Lower | 0.432 | 1.307 | 1.447 |
|  | Upper | 0.928 | 1.712 | 1.936 |
| **Chi squared** | | | χ^2^(3) = 110.752, p = 6.56 x10^-24^ | |
| **Nagelkerke R2 & AUC** | | | 0.129 & 68.1% | |
| **Classification predictive accuracy** | | | 74.0% | |
| **Sensitivity & Specificity** | | | 19.1% & 95.8% | |
| **Positive & Negative Predictive value** | | | 64.6% & 74.9% | |

**eTable 7.** Complete three-factor model results including the age-14 BOLD response to the anticipated ‘Large – small win’ condition in the right caudate.

| **N = 1178** | | **Right caudate BOLD**  **Age 14 Large – Small win**  **(-5.2 to 2.5)** | **EXT score**  **(-2.7 to 3.9)** | **CTQ**  **(-0.9 to 8.2)** |
| --- | --- | --- | --- | --- |
| Beta ± S.E. | | -0.253 (0.129) | 0.392 (0.07) | 0.511 (0.07) |
| Wald | | 3.858 | 32.664 | 47.201 |
| p | | 0.050 | 1.10 x10^-8^ | 6.41 x10^-12^ |
| Odds Ratio | | 0.776 | 1.481 | 1.666 |
| **95% CI for Odds Ratio** | Lower | 0.603 | 1.294 | 1.440 |
|  | Upper | 0.999 | 1.694 | 1.928 |
| **Chi squared** | | | χ^2^(3) = 109.077, p = 1.73 x10^-23^ | |
| **Nagelkerke R2 & AUC** | | | 0.127 & 68.3% | |
| **Classification predictive accuracy** | | | 74.4% | |
| **Sensitivity & Specificity** | | | 18.5% & 96.6% | |
| **Positive & Negative Predictive value** | | | 68.1% & 74.9% | |

**eTable 8.** Complete three-factor model results including the age-19 BOLD response to the anticipated ‘Large – no win’ condition in the right ventral striatum (VS).

| **N = 1174** | | **Right VS BOLD**  **Age 19 Large – No win**  **(-1.9 to 2.5)** | **EXT score**  **(-2.7 to 3.9)** | **CTQ**  **(-0.9 to 8.2)** |
| --- | --- | --- | --- | --- |
| Beta ± S.E. | | -0.310 (0.138) | 0.365 (0.07) | 0.519 (0.08) |
| Wald | | 5.025 | 27.923 | 46.618 |
| p | | 0.025 | 1.26 x10^-7^ | 8.626 x10^-12^ |
| Odds Ratio | | 0.734 | 1.440 | 1.681 |
| **95% CI for Odds Ratio** | Lower | 0.559 | 1.258 | 1.448 |
|  | Upper | 0.962 | 1.649 | 1.951 |
| **Chi squared** | | | χ^2^(3) = 101.630, p = 6.936 x10^-22^ | |
| **Nagelkerke R2 & AUC** | | | 0.118 & 67.1% | |
| **Classification predictive accuracy** | | | 73.5% | |
| **Sensitivity & Specificity** | | | 17.7% & 96.7% | |
| **Positive & Negative Predictive value** | | | 69.3% & 73.8% | |

*Complete model information for the four-factor models predicting one or more DSM-IV disorders at ages 14-19*

**eTable 9.** Complete four-factor model results including the age-14 BOLD response to the anticipated ‘Large – no win’ condition in the left ventral striatum (VS).

| **N=1145** | | **Left VS BOLD**  **Age 14 Large – No win**  **(-2.3 to 2.6)** | **EXT score**  **(-2.7 to 3.9)** | **CTQ**  **(-0.9 to 8.2)** | **Sex** |
| --- | --- | --- | --- | --- | --- |
| **Beta ± S.E.** | | -0.354 (0.15) | 0.414 (0.07) | 0.539 (0.08) | 0.798 (0.014) |
| **Wald** | | 5.369 | 33.892 | 47.550 | 30.133 |
| **p** | | 0.020 | 5.82 x10^-9^ | 5.36 x10^-12^ | 4.03 x10^-8^ |
| **Odds Ratio** | | 0.702 | 1.512 | 1.715 | 2.202 |
| **95% CI for Odds Ratio** | **Lower** | 0.520 | 1.316 | 1.471 | 1.661 |
|  | **Upper** | 0.947 | 1.738 | 1.998 | 2.919 |
| **Chi squared** | | | χ^2^(4) = 142.225, p = 9.42 x 10^-30^ | | |
| **Nagelkerke R^2^ & AUC** | | | 0.168 & 71.2% | | |
| **Classification predictive accuracy** | | | 74.7% | | |
| **Sensitivity & Specificity** | | | 22.8% & 95.2% | | |
| **Positive & Negative Predictive value** | | | 65.5% & 75.7% | | |

**eTable 10.** Complete four-factor model results including the age-14 BOLD response to the anticipated ‘Large – no win’ condition in the left caudate.

| **N = 1145** | | **Left caudate BOLD**  **Age 14 Large – No win**  **(-2.4 to 4.4)** | **EXT score**  **(-2.7 to 3.9)** | **CTQ**  **(-0.9 to 8.2)** | **Sex** |
| --- | --- | --- | --- | --- | --- |
| **Beta ± S.E.** | | -0.232 (0.111) | 0.410 (0.07) | 0.533 (0.08) | 0.779 (0.144) |
| **Wald** | | 4.399 | 33.232 | 46.570 | 29.348 |
| **p** | | 0.036 | 8.18 x10^-9^ | 8.84 x10^-12^ | 6.05 x10^-8^ |
| **Odds Ratio** | | 0.793 | 1.507 | 1.704 | 2.180 |
| **95% CI for Odds Ratio** | **Lower** | 0.638 | 1.311 | 1.462 | 1.644 |
|  | **Upper** | 0.985 | 1.732 | 1.986 | 2.890 |
| **Chi squared** | | | χ^2^(4) = 141.245 p = 1.59 x10^-29^ | | |
| **Nagelkerke R^2^ & AUC** | | | 0.167 & 71.5% | | |
| **Classification predictive accuracy** | | | 74.3% | | |
| **Sensitivity & Specificity** | | | 21.8% & 95.1% | | |
| **Positive & Negative Predictive value** | | | 64.0% & 75.4% | | |

**eTable 11.** Complete four-factor model results including the age-14 BOLD response to the anticipated ‘Large – no win’ condition in the right caudate.

| **N = 1145** | | **Right caudate BOLD**  **Age 14 Large – No win**  **(-2.2 to 3.1)** | **EXT score**  **(-2.7 to 3.9)** | **CTQ**  **(-0.9 to 8.2)** | **Sex** |
| --- | --- | --- | --- | --- | --- |
| **Beta ± S.E.** | | -0.254 (0.105) | 0.413 (0.07) | 0.530 (0.08) | 0.788 (0.144) |
| **Wald** | | 5.812 | 33.495 | 46.244 | 30.029 |
| **p** | | 0.016 | 7.14 x10^-9^ | 1.04 x10^-11^ | 4.26 x10^-8^ |
| **Odds Ratio** | | 0.776 | 1.511 | 1.698 | 2.199 |
| **95% CI for Odds Ratio** | **Lower** | 0.631 | 1.314 | 1.458 | 1.659 |
|  | **Upper** | 0.954 | 1.737 | 1.978 | 2.915 |
| **Chi squared** | | | χ^2^(4) = 142.683 p = 7.52 x10^-30^ | | |
| **Nagelkerke R^2^ & AUC** | | | 0.168 & 71.5% | | |
| **Classification predictive accuracy** | | | 74.8% | | |
| **Sensitivity & Specificity** | | | 23.4% & 95.2% | | |
| **Positive & Negative Predictive value** | | | 66.1% & 75.8% | | |

**eTable 12.** Complete four-factor model results including the age-14 BOLD response to the anticipated ‘Large – small win’ condition in the anterior cingulate cortex (ACC).

| **N = 1178** | | **ACC BOLD**  **Age 14 Large – Small win**  **(-3.6 to 3.5)** | **EXT score**  **(-2.7 to 3.9)** | **CTQ**  **(-0.9 to 8.2)** | **Sex** |
| --- | --- | --- | --- | --- | --- |
| **Beta ± S.E.** | | -0.362 (0.125) | 0.422 (0.07) | 0.532 (0.08) | 0.804 (0.142) |
| **Wald** | | 8.423 | 35.912 | 47.932 | 32.036 |
| **p** | | 0.004 | 2.06 x10^-9^ | 4.41 x10^-12^ | 1.51 x10^-8^ |
| **Odds Ratio** | | 0.696 | 1.525 | 1.703 | 2.235 |
| **95% CI for Odds Ratio** | **Lower** | 0.548 | 1.329 | 1.465 | 1.692 |
|  | **Upper** | 0.883 | 1.751 | 1.980 | 2.952 |
| **Chi squared** | | | χ^2^(4) = 146.389, p = 1.21 x10^-30^ | | |
| **Nagelkerke R^2^ & AUC** | | | 0.168 & 71.1% | | |
| **Classification predictive accuracy** | | | 75.2% | | |
| **Sensitivity & Specificity** | | | 24.2% & 95.5% | | |
| **Positive & Negative Predictive value** | | | 68.1% & 76.0% | | |

**eTable 13.** Complete four-factor model results including the age-14 BOLD response to the anticipated ‘Large – small win’ condition in the left putamen.

| **N = 1178** | | **Left putamen BOLD**  **Age 14 Large – Small win**  **(-2.0 to 3.9)** | **EXT score**  **(-2.7 to 3.9)** | **CTQ**  **(-0.9 to 8.2)** | **Sex** |
| --- | --- | --- | --- | --- | --- |
| **Beta ± S.E.** | | -0.261 (0.147) | 0.413 (0.07) | 0.532 (0.08) | 0.783 (0.142) |
| **Wald** | | 3.168 | 34.668 | 48.359 | 30.600 |
| **p** | | 0.075 | 3.91 x10^-9^ | 3.55 x10^-12^ | 3.17 x10^-8^ |
| **Odds Ratio** | | 0.770 | 1.511 | 1.703 | 2.188 |
| **95% CI for Odds Ratio** | **Lower** | 0.578 | 1.317 | 1.466 | 1.466 |
|  | **Upper** | 1.027 | 1.734 | 1.979 | 2.888 |
| **Chi squared** | | | χ^2^(4) = 141.079 p = 1.66 x10^-29^ | | |
| **Nagelkerke R^2^ & AUC** | | | 0.162 & 71.0% | | |
| **Classification predictive accuracy** | | | 74.3% | | |
| **Sensitivity & Specificity** | | | 22.1% & 95.0% | | |
| **Positive & Negative Predictive value** | | | 63.8% & 75.4% | | |

**eTable 14.** Complete four-factor model results including the age-14 BOLD response to the anticipated ‘Large – small win’ condition in the right ventral striatum.

| **N = 1178** | | **Right VS BOLD**  **Age 14 Large – Small win**  **(-1.7 to 2.2)** | **EXT score**  **(-2.7 to 3.9)** | **CTQ**  **(-0.9 to 8.2)** | **Sex** |
| --- | --- | --- | --- | --- | --- |
| **Beta ± S.E.** | | -0.429 (0.199) | 0.420 (0.07) | 0.532 (0.08) | 0.784 (0.142) |
| **Wald** | | 4.672 | 35.786 | 48.148 | 30.677 |
| **p** | | 0.031 | 2.20 x10^-9^ | 3.95 x10^-12^ | 3.05x10^-8^ |
| **Odds Ratio** | | 0.651 | 1.522 | 1.703 | 2.191 |
| **95% CI for Odds Ratio** | **Lower** | 0.441 | 1.326 | 1.465 | 1.660 |
|  | **Upper** | 0.961 | 1.746 | 1.979 | 2.892 |
| **Chi squared** | | | χ^2^(4) = 142.578 p = 7.92x10^-30^ | | |
| **Nagelkerke R^2^ & AUC** | | | 0.164 & 71.0% | | |
| **Classification predictive accuracy** | | | 74.7% | | |
| **Sensitivity & Specificity** | | | 22.4% & 95.5% | | |
| **Positive & Negative Predictive value** | | | 66.4% & 75.6% | | |

**eTable 15.** Complete four-factor model results including the age-14 BOLD response to the anticipated ‘Large – small win’ condition in the right caudate.

| **N = 1178** | | **Right caudate BOLD**  **Age 14 Large – Small win**  **(-5.2 to 2.5)** | **EXT score**  **(-2.7 to 3.9)** | **CTQ**  **(-0.9 to 8.2)** | **Sex** |
| --- | --- | --- | --- | --- | --- |
| **Beta ± S.E.** | | -0.265 (0.131) | 0.411 (0.07) | 0.528 (0.08) | 0.797 (0.142) |
| **Wald** | | 4.121 | 34.226 | 47.265 | 31.675 |
| **p** | | 0.042 | 4.91 x10^-9^ | 6.20 x10^-12^ | 1.82 x10^-8^ |
| **Odds Ratio** | | 0.767 | 1.509 | 1.696 | 2.219 |
| **95% CI for Odds Ratio** | **Lower** | 0.594 | 1.314 | 1.459 | 1.681 |
|  | **Upper** | 0.991 | 1.7301 | 1.971 | 2.929 |
| **Chi squared** | | | χ^2^(4) = 141.988 p = 1.06 x10^-29^ | | |
| **Nagelkerke R^2^ & AUC** | | | 0.163 & 71.2% | | |
| **Classification predictive accuracy** | | | 74.2% | | |
| **Sensitivity & Specificity** | | | 21.5% & 95.1% | | |
| **Positive & Negative Predictive value** | | | 63.7% & 75.3% | | |

**eTable 16.** Complete four-factor model results including the age-19 BOLD response to the anticipated ‘Large – no win’ condition in the right ventral striatum (VS).

| **N = 1174** | | **Right VS BOLD**  **Age 19 Large – No win**  **(-1.9 to 2.5)** | **EXT score**  **(-2.7 to 3.9)** | **CTQ**  **(-0.9 to 8.2)** | **Sex** |
| --- | --- | --- | --- | --- | --- |
| **Beta ± S.E.** | | -0.317 (0.140) | 0.385 (0.07) | 0.543 (0.08) | 0.718 (0.140) |
| **Wald** | | 5.110 | 29.686 | 48.483 | 26.671 |
| **p** | | 0.024 | 5.08 x10^-8^ | 3.33 x10^-12^ | 2.41 x10^-7^ |
| **Odds Ratio** | | 0.728 | 1.469 | 1.721 | 2.050 |
| **95% CI for Odds Ratio** | **Lower** | 0.553 | 1.279 | 1.477 | 1.561 |
|  | **Upper** | 0.959 | 1.687 | 2.005 | 2.692 |
| **Chi squared** | | | χ^2^(4) = 129.145, p = 5.93 x10^-27^ | | |
| **Nagelkerke R^2^ & AUC** | | | 0.148 & 69.2% | | |
| **Classification predictive accuracy** | | | 73.7% | | |
| **Sensitivity & Specificity** | | | 21.7% & 95.3% | | |
| **Positive & Negative Predictive value** | | | 65.8% & 74.5% | | |

*Complete model information for the three-factor models predicting DSM-IV and constructed SUDs*

**eTable 17.** Complete three-factor model results including the age-14 BOLD response to the anticipated ‘Large – no win’ condition in the left ventral striatum (VS) predicting both DSM-IV and constructed substance use disorders.

| **N = 1076** | | **Left VS BOLD**  **Age 14 Large – No win**  **(-2.3 to 2.6)** | **EXT score**  **(-2.7 to 3.9)** | **CTQ**  **(-0.9 to 8.2)** |
| --- | --- | --- | --- | --- |
| **Beta ± S.E.** | | -0.294 (0.15) | 0.463 (0.07) | 0.573 (0.08) |
| **Wald** | | 3.848 | 42.356 | 49.186 |
| **p** | | 0.050 | 7.60 x10^-11^ | 2.33 x10^-12^ |
| **Odds Ratio** | | 0.745 | 1.589 | 1.773 |
| **95% CI for Odds Ratio** | **Lower** | 0.555 | 1.382 | 1.511 |
|  | **Upper** | 1.000 | 1.827 | 2.080 |
| **Chi squared** | | | χ^2^(3) = 125.970, p = 3.99 x10^-27^ | |
| **Nagelkerke R2 & AUC** | | | 0.152 & 68.8% | |
| **Classification predictive accuracy** | | | 69.9% | |
| **Sensitivity & Specificity** | | | 28.9% & 92.0% | |
| **Positive & Negative Predictive value** | | | 66.1% & 70.6% | |

**eTable 18.** Complete three-factor model results including the age-14 BOLD response to the anticipated ‘Large – no win’ condition in the anterior cingulate cortex (ACC) predicting both DSM-IV and constructed substance use disorders.

| **N = 1076** | | **ACC BOLD**  **Age 14 Large – No win**  **(-2.3 to 2.6)** | **EXT score**  **(-2.7 to 3.9)** | **CTQ**  **(-0.9 to 8.2)** |
| --- | --- | --- | --- | --- |
| **Beta ± S.E.** | | -0.182 (0.10) | 0.471 (0.07) | 0.564 (0.08) |
| **Wald** | | 3.553 | 43.537 | 47.919 |
| **p** | | 0.059 | 4.16 x10^-11^ | 4.44 x10^-12^ |
| **Odds Ratio** | | 0.834 | 1.602 | 1.757 |
| **95% CI for Odds Ratio** | **Lower** | 0.690 | 1.393 | 1.498 |
|  | **Upper** | 1.007 | 1.842 | 2.061 |
| **Chi squared** | | | χ^2^(3) = 125.691, p = 4.59 x10^-27^ | |
| **Nagelkerke R2 & AUC** | | | 0.152 & 69.1% | |
| **Classification predictive accuracy** | | | 70.8% | |
| **Sensitivity & Specificity** | | | 30.8% & 92.4% | |
| **Positive & Negative Predictive value** | | | 68.6% & 71.2% | |

**eTable 19.** Complete three-factor model results including the age-14 BOLD response to the anticipated ‘Large – no win’ condition in the left caudate predicting both DSM-IV and constructed substance use disorders.

| **N = 1076** | | **Left caudate BOLD**  **Age 14 Large – No win**  **(-2.3 to 2.6)** | **EXT score**  **(-2.7 to 3.9)** | **CTQ**  **(-0.9 to 8.2)** |
| --- | --- | --- | --- | --- |
| **Beta ± S.E.** | | -0.206 (0.11) | 0.461 (0.07) | 0.564 (0.08) |
| **Wald** | | 3.612 | 41.947 | 47.840 |
| **p** | | 0.057 | 9.38 x10^-11^ | 4.63 x10^-12^ |
| **Odds Ratio** | | 0.814 | 1.586 | 1.758 |
| **95% CI for Odds Ratio** | **Lower** | 0.659 | 1.379 | 1.498 |
|  | **Upper** | 1.006 | 1.823 | 2.062 |
| **Chi squared** | | | χ^2^(3) = 125.738, p = 4.48 x10^-27^ | |
| **Nagelkerke R2 & AUC** | | | 0.152 & 69.0% | |
| **Classification predictive accuracy** | | | 70.4% | |
| **Sensitivity & Specificity** | | | 30.2% & 92.0% | |
| **Positive & Negative Predictive value** | | | 67.1% & 71.0% | |

**eTable 20.** Complete three-factor model results including the age-14 BOLD response to the anticipated ‘Large – no win’ condition in the right caudate predicting both DSM-IV and constructed substance use disorders.

| **N = 1076** | | **Right caudate BOLD**  **Age 14 Large – No win**  **(-2.2 to 3.1)** | **EXT score**  **(-2.7 to 3.9)** | **CTQ**  **(-0.9 to 8.2)** |
| --- | --- | --- | --- | --- |
| **Beta ± S.E.** | | -0.219 (0.10) | 0.464 (0.07) | 0.563 (0.08) |
| **Wald** | | 4.666 | 42.323 | 47.900 |
| **p** | | 0.031 | 7.74 x10^-11^ | 4.48 x10^-12^ |
| **Odds Ratio** | | 0.803 | 1.591 | 1.756 |
| **95% CI for Odds Ratio** | **Lower** | 0.659 | 1.383 | 1.497 |
|  | **Upper** | 0.980 | 1.830 | 2.060 |
| **Chi squared** | | | χ^2^(3) = 126.800, p = 2.65 x10^-27^ | |
| **Nagelkerke R2 & AUC** | | | 0.153 & 68.8% | |
| **Classification predictive accuracy** | | | 71.1% | |
| **Sensitivity & Specificity** | | | 31.0% & 92.7% | |
| **Positive & Negative Predictive value** | | | 69.6% & 71.4% | |

**eTable 21.** Complete three-factor model results including the age-14 BOLD response to the anticipated ‘Large – small win’ condition in the anterior cingulate cortex (ACC) predicting both DSM-IV and constructed substance use disorders.

| **N = 1105** | | **ACC BOLD**  **Age 14 Large – Small win**  **(-3.6 to 3.5)** | **EXT score**  **(-2.7 to 3.9)** | **CTQ**  **(-0.9 to 8.2)** |
| --- | --- | --- | --- | --- |
| **Beta ± S.E.** | | -0.243 (0.12) | 0.464 (0.07) | 0.557 (0.08) |
| **Wald** | | 4.007 | 43.408 | 48.656 |
| **p** | | ACC | 4.44 x10^-11^ | 3.05 x10^-12^ |
| **Odds Ratio** | | 0.784 | 1.590 | 1.745 |
| **95% CI for Odds Ratio** | **Lower** | 0.619 | 1.385 | 1.492 |
|  | **Upper** | 0.995 | 1.825 | 2.041 |
| **Chi squared** | | | χ^2^(3) = 125.929, p = 4.08 x10^-27^ | |
| **Nagelkerke R2 & AUC** | | | 0.148 & 68.7% | |
| **Classification predictive accuracy** | | | 70.5% | |
| **Sensitivity & Specificity** | | | 30.7% & 91.9% | |
| **Positive & Negative Predictive value** | | | 67.2% & 71.1% | |

**eTable 22.** Complete three-factor model results including the age-14 BOLD response to the anticipated ‘Large – small win’ condition in the midbrain predicting both DSM-IV and constructed substance use disorders.

| **N = 1105** | | **Midbrain BOLD**  **Age 14 Large – Small win**  **(-3.6 to 3.5)** | **EXT score**  **(-2.7 to 3.9)** | **CTQ**  **(-0.9 to 8.2)** |
| --- | --- | --- | --- | --- |
| **Beta ± S.E.** | | -0.427 (0.23) | 0.465 (0.07) | 0.554 (0.08) |
| **Wald** | | 3.608 | 43.438 | 48.699 |
| **p** | | 0.057 | 4.38 x10^-11^ | 2.99 x10^-12^ |
| **Odds Ratio** | | 0.652 | 1.592 | 1.739 |
| **95% CI for Odds Ratio** | **Lower** | 0.420 | 1.386 | 1.489 |
|  | **Upper** | 0.1.014 | 1.828 | 2.032 |
| **Chi squared** | | | χ^2^(3) = 125.567, p = 4.88 x10^-27^ | |
| **Nagelkerke R2 & AUC** | | | 0.148 & 68.4% | |
| **Classification predictive accuracy** | | | 70.2% | |
| **Sensitivity & Specificity** | | | 29.5% & 92.2% | |
| **Positive & Negative Predictive value** | | | 67.1 & 70.8% | |

**eTable 23.** Complete three-factor model results including the age-14 BOLD response to the anticipated ‘Large – small win’ condition in the right caudate predicting both DSM-IV and constructed substance use disorders.

| **N = 1105** | | **Right caudate BOLD**  **Age 14 Large – Small win**  **(-5.2 to 2.5)** | **EXT score**  **(-2.7 to 3.9)** | **CTQ**  **(-0.9 to 8.2)** |
| --- | --- | --- | --- | --- |
| **Beta ± S.E.** | | -0.266 (0.14) | 0.461 (0.07) | 0.554 (0.08) |
| **Wald** | | 3.818 | 42.988 | 48.050 |
| **p** | | 0.051 | 5.51 x10^-11^ | 4.15 x10^-12^ |
| **Odds Ratio** | | 0.766 | 1.586 | 1.740 |
| **95% CI for Odds Ratio** | **Lower** | 0.574 | 1.382 | 1.488 |
|  | **Upper** | 1.001 | 1.821 | 2.036 |
| **Chi squared** | | | χ^2^(3) = 125.737, p = 4.48 x10^-27^ | |
| **Nagelkerke R2 & AUC** | | | 0.148 & 68.8% | |
| **Classification predictive accuracy** | | | 70.3% | |
| **Sensitivity & Specificity** | | | 29.5% & 92.3% | |
| **Positive & Negative Predictive value** | | | 67.5% & 70.8% | |

**eTable 24.** Complete three-factor model results including the age-19 BOLD response to the anticipated ‘Large – no win’ condition in the right ventral striatum (VS) predicting both DSM-IV and constructed substance use disorders.

| **N = 1098** | | **Right VS BOLD**  **Age 19 Large – No win**  **(-1.9 to 2.5)** | **EXT score**  **(-2.7 to 3.9)** | **CTQ**  **(-0.9 to 8.2)** |
| --- | --- | --- | --- | --- |
| **Beta ± S.E.** | | -0.373 (0.14) | 0.433 (0.07) | 0.604 (0.08) |
| **Wald** | | 7.273 | 37.595 | 52.528 |
| **p** | | 0.007 | 8.71 x10^-10^ | 4.24 x10^-13^ |
| **Odds Ratio** | | 0.688 | 1.541 | 1.829 |
| **95% CI for Odds Ratio** | **Lower** | 0.525 | 1.342 | 1.554 |
|  | **Upper** | 0.903 | 1.770 | 2.154 |
| **Chi squared** | | | χ^2^(3) = 126.431, p = 3.18 x10^-27^ | |
| **Nagelkerke R2 & AUC** | | | 0.149 & 68.8% | |
| **Classification predictive accuracy** | | | 69.5% | |
| **Sensitivity & Specificity** | | | 30.7% & 91.2% | |
| **Positive & Negative Predictive value** | | | 66.1% & 70.2% | |

**eTable 25.** Complete three-factor model results including the age-19 BOLD response to the anticipated ‘Large – no win’ condition in the right caudate predicting both DSM-IV and constructed substance use disorders.

| **N = 1098** | | **Right caudate BOLD**  **Age 19 Large – No win**  **(-2.4 to 2.9)** | **EXT score**  **(-2.7 to 3.9)** | **CTQ**  **(-0.9 to 8.2)** |
| --- | --- | --- | --- | --- |
| **Beta ± S.E.** | | -0.243 (0.10) | 0.433 (0.07) | 0.605 (0.08) |
| **Wald** | | 6.094 | 37.686 | 52.649 |
| **p** | | 0.014 | 8.31 x10^-10^ | 3.98 x10^-13^ |
| **Odds Ratio** | | 0.784 | 1.542 | 1.832 |
| **95% CI for Odds Ratio** | **Lower** | 0.647 | 1.343 | 1.555 |
|  | **Upper** | 0.951 | 1.770 | 2.156 |
| **Chi squared** | | | χ^2^(3) = 125.179, p = 5.91 x10^-27^ | |
| **Nagelkerke R2 & AUC** | | | 0.148 & 68.5% | |
| **Classification predictive accuracy** | | | 68.9% | |
| **Sensitivity & Specificity** | | | 29.4% & 90.9% | |
| **Positive & Negative Predictive value** | | | 64.4% & 69.7% | |

**eTable 26.** Complete three-factor model results including the age-19 BOLD response to the anticipated ‘Small – no win’ condition in the left caudate predicting both DSM-IV and constructed substance use disorders.

| **N = 1097** | | **Left Caudate BOLD**  **Age 19 Small – No win**  **(-2.0 to 2.2)** | **EXT score**  **(-2.7 to 3.9)** | **CTQ**  **(-0.9 to 8.2)** |
| --- | --- | --- | --- | --- |
| **Beta ± S.E.** | | -0.265 (0.12) | 0.424 (0.07) | 0.604 (0.08) |
| **Wald** | | 5.256 | 36.404 | 52.846 |
| **p** | | 0.022 | 1.60 x10^-9^ | 3.60 x10^-13^ |
| **Odds Ratio** | | 0.767 | 1.528 | 1.830 |
| **95% CI for Odds Ratio** | **Lower** | 0.612 | 1.332 | 1.555 |
|  | **Upper** | 0.962 | 1.754 | 2.153 |
| **Chi squared** | | | χ^2^(3) = 124.689, p = 7.54 x10^-27^ | |
| **Nagelkerke R2 & AUC** | | | 0.147 & 68.3% | |
| **Classification predictive accuracy** | | | 69.8% | |
| **Sensitivity & Specificity** | | | 31.5% & 91.3% | |
| **Positive & Negative Predictive value** | | | 67.0% & 70.4% | |

*Effect of Sex in SUD models*

For each model where the fMRI contrast was a significant predictor in the models including SUD diagnoses, sex was tested as a fourth factor. In each of these models, sex was a significant predictor (*p* < 0.005). As with the DSM-IV only models, being female slightly increased the odds of being classified as having a disorder (OR ≈ 1.400 (95% CI = 1.077 – 1.823)), though the effect was weaker.

**eTable 27.** Complete four-factor model results including the age-14 BOLD response to the anticipated ‘Large – no win’ condition in the left ventral striatum (VS) predicting both DSM-IV and constructed substance use disorders.

| **N = 1076** | | **Left VS BOLD**  **Age 14 Large – No win**  **(-2.3 to 2.6)** | **EXT score**  **(-2.7 to 3.9)** | **CTQ**  **(-0.9 to 8.2)** | **Sex** |
| --- | --- | --- | --- | --- | --- |
| **Beta ± S.E.** | | -0.291 (0.15) | 0.465 (0.07) | 0.583 (0.08) | 0.446 (0.14) |
| **Wald** | | 3.723 | 42.108 | 49.314 | 10.486 |
| **p** | | 0.054 | 8.64 x10^-11^ | 2.18 x10^-12^ | 0.001 |
| **Odds Ratio** | | 0.748 | 1.592 | 1.791 | 1.563 |
| **95% CI for Odds Ratio** | **Lower** | 0.556 | 1.384 | 1.522 | 1.193 |
|  | **Upper** | 1.005 | 1.833 | 2.107 | 2.048 |
| **Chi squared** | | | χ^2^(4) = 136.579, p = 1.52 x10^-28^ | | |
| **Nagelkerke R2 & AUC** | | | 0.164 & 70.0% | | |
| **Classification predictive accuracy** | | | 70.0% | | |
| **Sensitivity & Specificity** | | | 30.2% & 91.4% | | |
| **Positive & Negative Predictive value** | | | 65.5% & 70.8% | | |

**eTable 28.** Complete four-factor model results including the age-14 BOLD response to the anticipated ‘Large – no win’ condition in the anterior cingulate cortex (ACC) predicting both DSM-IV and constructed substance use disorders.

| **N = 1076** | | **ACC BOLD**  **Age 14 Large – No win**  **(-2.3 to 2.6)** | **EXT score**  **(-2.7 to 3.9)** | **CTQ**  **(-0.9 to 8.2)** | **Sex** |
| --- | --- | --- | --- | --- | --- |
| **Beta ± S.E.** | | -0.170 (0.10) | 0.473 (0.07) | 0.574 (0.08) | 0.438 (0.14) |
| **Wald** | | 3.042 | 43.350 | 48.031 | 10.099 |
| **p** | | 0.081 | 4.58 x10^-11^ | 4.20 x10^-12^ | 0.001 |
| **Odds Ratio** | | 0.844 | 1.605 | 1.775 | 1.550 |
| **95% CI for Odds Ratio** | **Lower** | 0.697 | 1.394 | 1.509 | 1.183 |
|  | **Upper** | 1.021 | 1.848 | 2.088 | 2.031 |
| **Chi squared** | | | χ^2^(4) = 135.904, p = 2.13 x10^-28^ | | |
| **Nagelkerke R2 & AUC** | | | 0.163 & 70.0% | | |
| **Classification predictive accuracy** | | | 70.1% | | |
| **Sensitivity & Specificity** | | | 31.3% & 91.0% | | |
| **Positive & Negative Predictive value** | | | 65.2% & 71.1% | | |

**eTable 29.** Complete four-factor model results including the age-14 BOLD response to the anticipated ‘Large – no win’ condition in the left caudate predicting both DSM-IV and constructed substance use disorders.

| **N = 1076** | | **Left caudate BOLD**  **Age 14 Large – No win**  **(-2.3 to 2.6)** | **EXT score**  **(-2.7 to 3.9)** | **CTQ**  **(-0.9 to 8.2)** | **Sex** |
| --- | --- | --- | --- | --- | --- |
| **Beta ± S.E.** | | -0.191 (0.11) | 0.463 (0.07) | 0.574 (0.08) | 0.438 (0.14) |
| **Wald** | | 3.064 | 41.787 | 48.049 | 10.064 |
| **p** | | 0.080 | 1.02 x10^-10^ | 4.16 x10^-12^ | 0.002 |
| **Odds Ratio** | | 0.826 | 1.590 | 1.776 | 1.549 |
| **95% CI for Odds Ratio** | **Lower** | 0.667 | 1.381 | 1.510 | 1.182 |
|  | **Upper** | 1.023 | 1.829 | 2.089 | 2.030 |
| **Chi squared** | | | χ^2^(4) = 135.915, p = 2.11 x10^-28^ | | |
| **Nagelkerke R2 & AUC** | | | 0.163 & 70.0% | | |
| **Classification predictive accuracy** | | | 70.8% | | |
| **Sensitivity & Specificity** | | | 31.3% & 92.1% | | |
| **Positive & Negative Predictive value** | | | 68.2% & 71.3% | | |

**eTable 30.** Complete four-factor model results including the age-14 BOLD response to the anticipated ‘Large – no win’ condition in the right caudate predicting both DSM-IV and constructed substance use disorders.

| **N = 1076** | | **Right caudate BOLD**  **Age 14 Large – No win**  **(-2.3 to 2.6)** | **EXT score**  **(-2.7 to 3.9)** | **CTQ**  **(-0.9 to 8.2)** | **Sex** |
| --- | --- | --- | --- | --- | --- |
| **Beta ± S.E.** | | -0.218 (0.1) | 0.466 (0.07) | 0.573 (0.08) | 0.446 (0.14) |
| **Wald** | | 4.530 | 42.076 | 47.896 | 10.477 |
| **p** | | 0.033 | 8.78 x10^-11^ | 4.49 x10^-12^ | 0.001 |
| **Odds Ratio** | | 0.804 | 1.594 | 1.773 | 1.563 |
| **95% CI for Odds Ratio** | **Lower** | 0.658 | 1.385 | 1.508 | 1.193 |
|  | **Upper** | 0.983 | 1.836 | 2.085 | 2.048 |
| **Chi squared** | | | χ^2^(4) = 137.399, p = 1.02 x10^-28^ | | |
| **Nagelkerke R2 & AUC** | | | 0.165 & 70.1% | | |
| **Classification predictive accuracy** | | | 70.6% | | |
| **Sensitivity & Specificity** | | | 31.6% & 91.7% | | |
| **Positive & Negative Predictive value** | | | 67.2% & 71.3% | | |

**eTable 31.** Complete four-factor model results including the age-14 BOLD response to the anticipated ‘Large – small win’ condition in the anterior cingulate cortex (ACC) predicting both DSM-IV and constructed substance use disorders.

| **N = 1105** | | **ACC BOLD**  **Age 14 Large – Small win**  **(-3.6 to 3.5)** | **EXT score**  **(-2.7 to 3.9)** | **CTQ**  **(-0.9 to 8.2)** | **Sex** |
| --- | --- | --- | --- | --- | --- |
| **Beta ± S.E.** | | -0.253 (0.12) | 0.437 (0.07) | 0.516 (0.08) | 0.449 (0.14) |
| **Wald** | | 4.290 | 36.367 | 48.578 | 10.785 |
| **p** | | 0.038 | 3.34 x10^-11^ | 3.03 x10^-12^ | 7.96 x10^-4^ |
| **Odds Ratio** | | 0.777 | 1.595 | 1.761 | 1.578 |
| **95% CI for Odds Ratio** | **Lower** | 0.611 | 1.388 | 1.502 | 1.209 |
|  | **Upper** | 0.987 | 1.833 | 2.065 | 2.061 |
| **Chi squared** | | | χ^2^(3) = 137.321, p = 1.06 x10^-28^ | | |
| **Nagelkerke R2 & AUC** | | | 0.161 & 69.7% | | |
| **Classification predictive accuracy** | | | 70.3% | | |
| **Sensitivity & Specificity** | | | 31.0% & 91.5% | | |
| **Positive & Negative Predictive value** | | | 66.3% & 71.1% | | |

**eTable 32.** Complete four-factor model results including the age-14 BOLD response to the anticipated ‘Large – small win’ condition in the midbrain predicting both DSM-IV and constructed substance use disorders.

| **N = 1105** | | **Midbrain BOLD**  **Age 14 Large – Small win**  **(-3.6 to 3.5)** | **EXT score**  **(-2.7 to 3.9)** | **CTQ**  **(-0.9 to 8.2)** | **Sex** |
| --- | --- | --- | --- | --- | --- |
| **Beta ± S.E.** | | -0.468 (0.23) | 0.468 (0.07) | 0.562 (0.08) | 0.465 (0.14) |
| **Wald** | | 4.276 | 43.392 | 48.617 | 11.650 |
| **p** | | 0.039 | 4.48 x10^-11^ | 3.11 x10^-12^ | 6.42 x10^-4^ |
| **Odds Ratio** | | 0.626 | 1.597 | 1.754 | 1.592 |
| **95% CI for Odds Ratio** | **Lower** | 0.402 | 1.389 | 1.498 | 1.219 |
|  | **Upper** | 0.976 | 1.836 | 2.055 | 2.079 |
| **Chi squared** | | | χ^2^(3) = 137.371, p = 1.03 x10^-28^ | | |
| **Nagelkerke R2 & AUC** | | | 0.161 & 69.7% | | |
| **Classification predictive accuracy** | | | 70.0% | | |
| **Sensitivity & Specificity** | | | 30.0% & 91.6% | | |
| **Positive & Negative Predictive value** | | | 65.9% & 70.8% | | |

**eTable 33.** Complete four-factor model results including the age-14 BOLD response to the anticipated ‘Large – small win’ condition in the right caudate predicting both DSM-IV and constructed substance use disorders.

| **N = 1105** | | **Right caudate BOLD**  **Age 14 Large – Small win**  **(-5.2 to 2.5)** | **EXT score**  **(-2.7 to 3.9)** | **CTQ**  **(-0.9 to 8.2)** | **Sex** |
| --- | --- | --- | --- | --- | --- |
| **Beta ± S.E.** | | -0.270 (0.14) | 0.465 (0.07) | 0.563 (0.08) | 0.452 (0.14) |
| **Wald** | | 3.875 | 42.903 | 48.062 | 11.037 |
| **p** | | 0.049 | 5.75 x10^-11^ | 4.13 x10^-12^ | 8.93 x10^-4^ |
| **Odds Ratio** | | 0.764 | 1.591 | 1.756 | 1.571 |
| **95% CI for Odds Ratio** | **Lower** | 0.584 | 1.385 | 1.498 | 1.203 |
|  | **Upper** | 0.999 | 1.829 | 2.060 | 2.051 |
| **Chi squared** | | | χ^2^(4) = 136.909, p = 1.29 x10^-28^ | | |
| **Nagelkerke R2 & AUC** | | | 0.160 & 69.8% | | |
| **Classification predictive accuracy** | | | 70.0% | | |
| **Sensitivity & Specificity** | | | 30.5% & 91.2% | | |
| **Positive & Negative Predictive value** | | | 65.2% & 70.9% | | |

**eTable 34.** Complete four-factor model results including the age-19 BOLD response to the anticipated ‘Large – no win’ condition in the right ventral striatum (VS) predicting both DSM-IV and constructed substance use disorders.

| **N = 1098** | | **Right VS BOLD**  **Age 19 Large – No win**  **(-1.9 to 2.5)** | **EXT score**  **(-2.7 to 3.9)** | **CTQ**  **(-0.9 to 8.2)** | **Sex** |
| --- | --- | --- | --- | --- | --- |
| **Beta ± S.E.** | | -0.373 (0.14) | 0.437 (0.07) | 0.618 (0.08) | 0.414 (0.14) |
| **Wald** | | 7.214 | 37.779 | 53.715 | 9.319 |
| **p** | | 0.007 | 7.92 x10^-10^ | 2.32 x10^-13^ | 0.002 |
| **Odds Ratio** | | 0.688 | 1.548 | 1.856 | 1.513 |
| **95% CI for Odds Ratio** | **Lower** | 0.524 | 1.347 | 1.573 | 1.160 |
|  | **Upper** | 0.904 | 1.780 | 2.189 | 1.973 |
| **Chi squared** | | | χ^2^(4) = 135.851 p = 2.18 x10^-28^ | | |
| **Nagelkerke R2 & AUC** | | | 0.160 & 69.8% | | |
| **Classification predictive accuracy** | | | 69.2% | | |
| **Sensitivity & Specificity** | | | 30.5% & 90.9% | | |
| **Positive & Negative Predictive value** | | | 65.2% & 70.0% | | |

**eTable 35.** Complete four-factor model results including the age-19 BOLD response to the anticipated ‘Large – no win’ condition in the right caudate both DSM-IV predicting and constructed substance use disorders.

| **N = 1098** | | **Right caudate BOLD**  **Age 19 Large – No win**  **(-2.4 to 2.9)** | **EXT score**  **(-2.7 to 3.9)** | **CTQ**  **(-0.9 to 8.2)** | **Sex** |
| --- | --- | --- | --- | --- | --- |
| **Beta ± S.E.** | | -0.228 (0.10) | 0.416 (0.07) | 0.613 (0.08) | 0.400 (0.14) |
| **Wald** | | 5.324 | 32.807 | 53.137 | 8.628 |
| **p** | | 0.021 | 7.82 x10^-10^ | 2.32 x10^-13^ | 0.003 |
| **Odds Ratio** | | 0.796 | 1.548 | 1.855 | 1.489 |
| **95% CI for Odds Ratio** | **Lower** | 0.656 | 1.347 | 1.573 | 1.141 |
|  | **Upper** | 0.966 | 1.779 | 2.189 | 1.942 |
| **Chi squared** | | | χ^2^(4) = 133.878, p = 5.78 x10^-28^ | | |
| **Nagelkerke R2 & AUC** | | | 0.157 & 69.5% | | |
| **Classification predictive accuracy** | | | 68.9% | | |
| **Sensitivity & Specificity** | | | 29.4% & 90.9% | | |
| **Positive & Negative Predictive value** | | | 64.4% & 69.7% | | |

**eTable 36.** Complete four-factor model results including the age-19 BOLD response to the anticipated ‘Small – no win’ condition in the left caudate predicting both DSM-IV and constructed substance use disorders.

| **N = 1097** | | **Left caudate BOLD**  **Age 19 Small – No win**  **(-2.4 to 2.9)** | **EXT score**  **(-2.7 to 3.9)** | **CTQ**  **(-0.9 to 8.2)** | **Sex** |
| --- | --- | --- | --- | --- | --- |
| **Beta ± S.E.** | | -0.248 (0.12) | 0.427 (0.07) | 0.617 (0.08) | 0.389 (0.14) |
| **Wald** | | 4.529 | 36.350 | 53.826 | 8.251 |
| **p** | | 0.033 | 1.65 x10^-9^ | 2.19 x10^-13^ | 0.004 |
| **Odds Ratio** | | 0.780 | 1.532 | 1.853 | 1.475 |
| **95% CI for Odds Ratio** | **Lower** | 0.621 | 1.334 | 1.572 | 1.132 |
|  | **Upper** | 0.981 | 1.760 | 2.185 | 1.924 |
| **Chi squared** | | | χ^2^(4) = 133.016, p = 8.81 x10^-28^ | | |
| **Nagelkerke R2 & AUC** | | | 0.157 & 69.1% | | |
| **Classification predictive accuracy** | | | 69.7% | | |
| **Sensitivity & Specificity** | | | 31.0% & 91.5% | | |
| **Positive & Negative Predictive value** | | | 67.0% & 70.3% | | |

*Anhedonia and Ventral Striatum*

Stringaris and colleagues (2015; 7) reported that anhedonia is associated with reduced MID task-induced ventral striatal (VS) activations in depressed adolescents. To determine whether anhedonia was associated with VS BOLD responses in our sample, we conducted a point-biserial correlation between striatal BOLD signal and the loss of interest item from the DAWBA depression screening (‘In the last 4 weeks, have there been times when you have lost interest in everything, or nearly everything, that you normally enjoy doing?’). Individuals were coded as ‘1’ if they responded yes to this item, and ‘0’ if they responded no. A random sample of individuals who did not endorse anhedonia was selected to create balanced groups. A weak effect was observed in the VS; low responses in the left VS were associated with higher anhedonia (Lg – No win; *r*_PB_ = r = -0.089, p = 0.044, one-tailed).
